# Exploring how PRIME-Parkinson care is implemented and whether, how and why it produces change, for who and under what conditions: a protocol for an embedded process evaluation within the PRIME-UK randomised controlled trial

**DOI:** 10.1101/2024.12.23.24319541

**Authors:** Katherine Lloyd, Emma Tenison, Safi Smith, Fiona Lithander, Judi Kidger, Heather Brant, Sabi Redwood, Yoav Ben-Shlomo, Emily J Henderson

## Abstract

**Introduction:** The PRIME-UK randomised controlled trial (RCT) aims to establish whether a model of care that seeks to be proactive, integrated and empower participants, caregivers and healthcare professionals can improve outcomes in people with parkinsonism. Given that this intervention is novel and complex, understanding whether and how the intervention will be acceptable, implementable, cost effective and scalable across contexts are key questions beyond that of whether ‘it works’. We describe an embedded process evaluation to answer these questions, which aims to support interpretation of the trial results, refinement of the intervention and support future scaling of the PRIME-Parkinson model of care.

**Methods and analysis:** A mixed methods approach will be used to collect data across four process evaluation domains: implementation, mechanism of change, acceptability and context. Quantitative data will be collected prospectively from all participants and analysed descriptively with exploratory tests of relationships as power allows. Qualitative data will be collected through semi-structured interviews with a purposively sampled sub-population of participants, caregivers and staff members as well as case studies where relevant. Interview transcripts will be analysed thematically using interpretive qualitative analysis. Synthesis of quantitative and qualitative data will also be performed to draw conclusions.

**Ethics and dissemination:** The quantitative data will be collected as part of the main PRIME-UK RCT which was been granted NHS REC approval (21/LO/0387) on 27th July 2021. The qualitative data will be collected as part of a sub-study, ‘PRIME-Qual’, which was granted NHS REC approval (21/LO/0388) on 14th July 2021. The mixed methods process evaluation will be published after the conclusion of the trial in addition to the main trial findings.

**Trial registration number:** NCT05127057

**Data availability statement:** Access to the data will be available through application to the Chief Investigator. Pseudo-anonymised data may be shared with other researchers to enable knowledge synthesis.

**ARTICLE SUMMARY:** *Strengths and limitations of this study:*

- This is a pre-planned process evaluation of a complex intervention, which uses robust mixed methodology to support understanding and interpretation of the main trial findings
- This process evaluation is embedded within a RCT which aims to recruit a representative sample of people living with parkinsonism, meaning the process evaluation has potential to gather a broad range of perspectives
- Our mixed methods approach provides breadth of data across the study population and more in-depth exploration of themes in a purposively sampled sub-population
- The main limitation is that statistical power will limit the use of statistical tests in the analysis of quantitative data across subgroups.

## INTRODUCTION

### The PRIME-UK Randomised Controlled Trial

Parkinsonism is a clinical syndrome which typically causes motor symptoms including tremor, bradykinesia, rigidity and postural instability as well as a range of non-motor symptoms. The most common cause is idiopathic Parkinson’s disease (PD) but the syndrome of parkinsonism also encompasses other conditions such as Dementia with Lewy Bodies (DLB), Progressive Supranuclear Palsy (PSP), Multiple System Atrophy (MSA) and Corticobasal Degeneration (CBD). There is a high degree of heterogeneity of symptoms, response to treatment, prognosis and therefore the experience of having parkinsonism varies widely between individuals.

More than 6 million people are affected worldwide [1] making PD the fastest growing neurological condition;[2] the prevalence has more than doubled in the last 25 years, predominantly due to the ageing population worldwide, and projections predict a similar rate of increase over the next 30.[1] PD reduces life expectancy[3] and quality of life with increasing effect as the disease progresses[4] and, with this rising prevalence, the global burden of PD measured using disability-adjusted life years (DALYs) is also increasing.[2] This impact extends to friends and family who commonly act as informal caregivers.[5] The direct healthcare-associated costs for people with PD are high,[6] and the increasing prevalence creates economic implications for health care systems.

The high and rising global burden of PD means that good quality healthcare is required to maximise positive outcomes for people with PD, whilst reducing wasted time and resources.[7] The World Health Organisation describe good quality healthcare as “the right care, at the right time, in a coordinated way, responding to the service users’ needs and preferences, while minimising harm and resource waste”.[7] There are recognised inadequacies in the current healthcare delivery in PD compromising its quality. PD care lacks personalisation, often taking a ‘one-size fits all’ approach despite recognised heterogeneity in those affected,[8] and certain aspects of care are frequently overlooked, such as non-motor symptoms and bone health.[9,10] The approach to care is often reactive in nature, for example people with PD are only referred for falls prevention interventions after their first fall.[11] Owing to the fragmentation in care provided and the lack of coordination between healthcare providers, people with PD urgently need a single point of access to help and support, as well as increased empowerment to self-manage their condition.[12]

Based on this consensus regarding shortcomings in current care[13] we proposed a new model of care termed PRIME-Parkinson care for people with parkinsonism.[14] This model is designed to provide personalised and integrated care, manage problems proactively and empower patients and caregivers. To gather evidence on its effect on health-related outcomes and quality of life, two studies are ongoing. In the Netherlands, an observational study is introducing the PRIME-NL model of care in an index region and comparing outcomes to other ‘usual care’ regions.[15] In the UK, the PRIME-UK RCT, is a single centre study which has recruited 214 people with parkinsonism (excluding drug induced PD) and randomised them to receive ‘usual care’ or ‘PRIME-Parkinson care’ and ‘usual care’. Caregivers are also given the option to enrol in a sub-study. The new care model intervention will be delivered for 24 months, at which point the primary outcome of goal attainment and other secondary measures of health and wellbeing will be captured.[16] Full details relating to the recruitment, design, delivery and outcome assessment can be found in the published protocol.[16]

### The role of process evaluation within the PRIME-RCT

The PRIME-Parkinson model of care is a complex, multi-component intervention acting on multiple domains of health and wellbeing, which is delivered flexibly depending on participant need and which acts within an existing healthcare system. [16] The proposed causal effect of the intervention is hypothetical and multi-staged. Research involving complex interventions, such as this, moves away from answering a “binary question of effectiveness” but rather aims to establish “whether and how the intervention will be acceptable, implementable, cost effective and scalable across contexts”.[17,18] The perspective of research changes to a theory and systems approach, understanding how and in which circumstances change is brought about and interacts with the system in which it is introduced.[18] This process supports us to have greater confidence in the conclusions drawn from studies of effectiveness.[18]

To answer these broader questions regarding the PRIME-Parkinson intervention in the UK, we will conduct an embedded process evaluation, with relevant process evaluation outcome measure data collected in parallel with the trial effectiveness data. The protocol for this embedded process evaluation is outlined in this paper. The potential merits of a process evaluation within the PRIME-UK RCT are four-fold: 1) to guide refinement of the intervention iteratively throughout the study in response to data accumulated on implementation quantity, quality, reach and acceptability, 2) to refine the intervention theory through generating knowledge of the mechanisms of change activated, 3) to support interpretation of the study outcomes as to why it improved/failed to improve outcomes, 4) to allow scalability of the intervention into other settings and into a ‘real-world’ context. This final point is of particular importance; the effect of PRIME-Parkinson intervention will purely show that it was beneficial, or not, to the population that received it in that setting, which is removed from improving outcomes for people with parkinsonism across the UK.

Given the recognised heterogeneity of both PD care services in the UK[10] and the experiences of living with PD, understanding the context around the implementation of the PRIME-UK RCT will be invaluable in future multi-centre studies and for implementation into routine care. The process evaluation allows us to understand the impact of the intervention dependent on patient and service level characteristics. Scaling the project based on effectiveness across this study population alone risks missing the nuance of for whom the intervention is effective. Given the concerted effort in the PRIME-UK RCT to reach underserved groups, we have a duty to consider the intervention and outcomes from their perspectives.[19]

### Aims and objectives

The overarching aim of this process evaluation is to capture how the PRIME-Parkinson intervention is implemented and whether, how and why it produces change, for who and under what conditions. This can influence refinement of the intervention iteratively throughout the study in response to data accumulated, develop our understanding of the theory of change and support the implementation of the PRIME-Parkinson model of care into other settings or into ‘real-world practice’.

The objectives are to:

1. Assess how successfully the intervention is being **implemented** (quantity, quality, reach, engagement), whether this shows **fidelity** with the overarching aims of the PRIME intervention
2. Establish whether the **mechanisms of change,** in relation to the logic model, are being activated
3. Explore the **acceptability** of the intervention from the viewpoint of the participants and caregivers, including factors that shape experience of taking part and adherence to intervention
4. For each of the above domains, examine how **context**, such as participant and service level factors affect implementation, outcomes, acceptability and mechanisms of change of the PRIME intervention

Our approach to the process evaluation with the PRIME-UK RCT will use the theory perspective as described in the Medical Research Council (MRC) guidance.[18] Within this approach there is less distinction between process and effectiveness outcomes and instead the focus is on understanding the impact of different components to generate knowledge and develop our understanding of how we should be intervening to improve outcomes in people with PD. The relevance and merits of the four proposed process evaluation domains are outlined below.

### Components of the Process Evaluation

#### Intervention implementation

Implementation aims to capture what, regarding the PRIME-Parkinsonism intervention, is delivered in practice and to whom.[18] This can support interpretation of the study outcomes by understanding ‘what works’ and is a required component for future research or translation into clinical care. We will collect data from multiple different sources to capture (1) the quantity and quality of the intervention delivered, (2) the fidelity of the intervention – how it relates to the intervention described in the protocol and whether the overall ethos of the PRIME-Parkinson intervention was upheld, (3) the reach of the intervention – who received and engaged with the different components of the intervention.

#### Mechanisms of change

The proposed mechanisms of change for how the PRIME-Parkinson model of care may augment an individual’s ability to achieve their personal goals, and positively impact health and wellbeing are highlighted in the logic model (see figure 1). This logic model represents an iteration of the PRIME logic models, which used 6 recognised “problems with current care” to drive intervention development by showing how potential strategies and activities could produce change towards desired outcomes. [14] The new logic model takes the perspective of PRIME-Parkinson as a composite intervention, which we are now testing for efficacy in a complex RCT. It focuses on how the intervention is hypothesised to bring about change, which may be an effect of individual components, the interaction of multiple components, as well as the interaction with the context within which the intervention is implemented. The proposed mechanisms have been synthesised from a variety of sources including the existing literature, expert opinions, and the perspectives and experiences of the public and patients. This domain of the process evaluation aims to test hypothesised causal pathways defined in the logic model using mixed quantitative and qualitative methods. This process will further our understanding of the PRIME-Parkinson programme theory.

**Figure 1.**
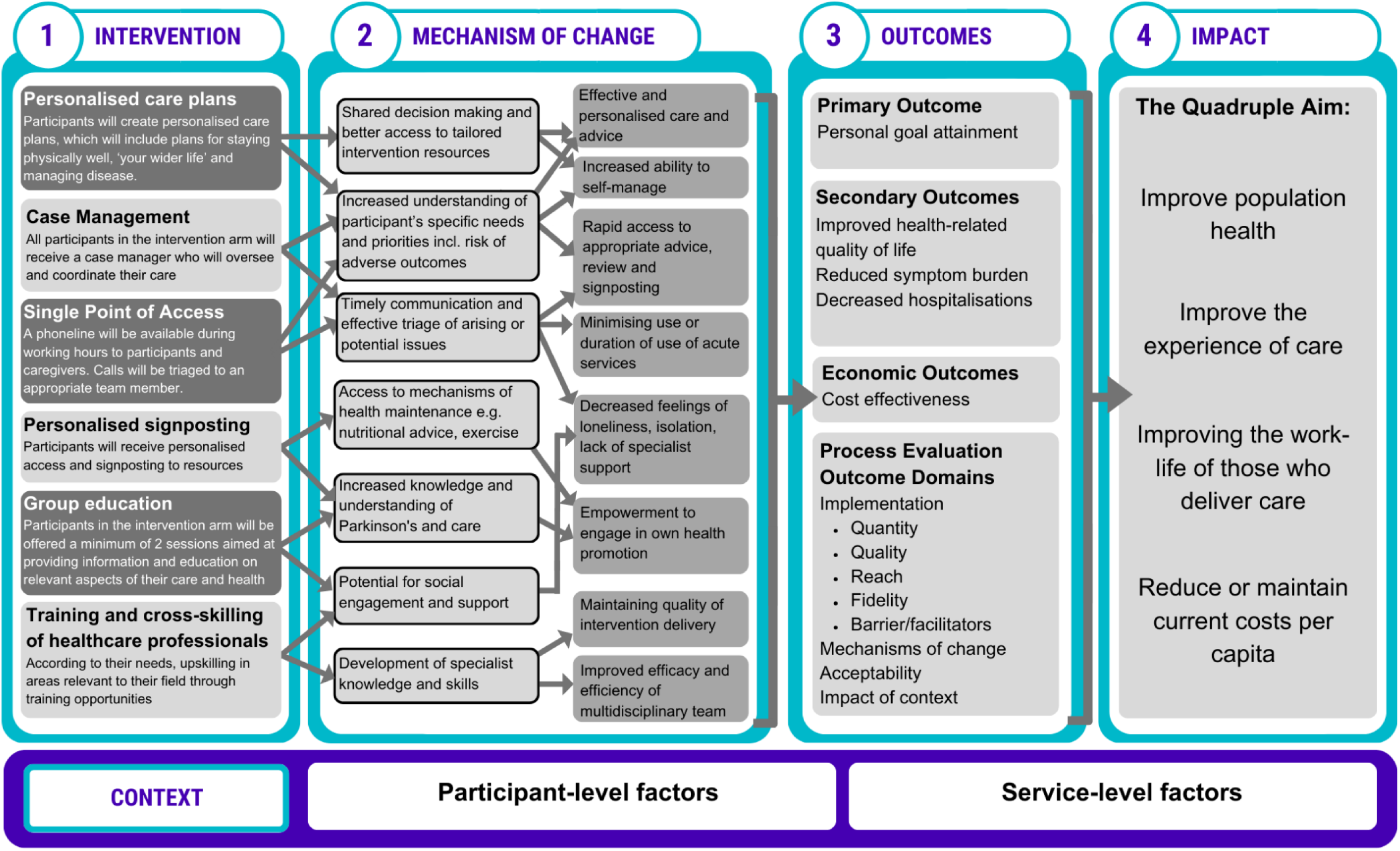
Logic model of the PRIME intervention showing components, hypothesised mechanisms of change to produce study outcomes and real world impact

#### Acceptability

Given this is the first evaluation of the PRIME-Parkinson intervention, we will also explore the acceptability of the intervention from the perspective of participants, caregivers and healthcare professionals through assessing their experiences.

#### Context

The PRIME-Parkinson model of care is being implemented within an existing system, with which it will interact.[18] Service level factors such as the existing infrastructure of Parkinson’s care, the components of which are known to differ across UK services[10] and locally between areas, may influence how PRIME-Parkinson care is implemented, produces change and generates outcomes. Participant level factors such as epidemiological characteristics including disease specific measures (diagnosis, stage, presence of cognitive impairment, relationship with current consultant, readiness of access to care), socio-economic factors (having a caregiver, social support network, index of multiple deprivation) and geographical factors (urban versus rural participants, access to transport) may also have a role. The impact of external shocks, such as changes to care post the Covid-19 pandemic, will also be considered. For example, whether a depleted ‘usual care’, which may not be representative of ‘normal’, impacted the trial. Capturing these contextual factors complements the previously described domains of the process evaluation. We will evaluate whether contextual factors impact how the intervention was implemented, the mechanisms of change, acceptability and observed outcomes. Understanding the impact of contextual factors allows us to consider how the intervention may need to be refined to serve certain groups and settings.

The research questions, within each of the above domains, addressed by the process evaluation are outlined in table 1.

**Table 1.**
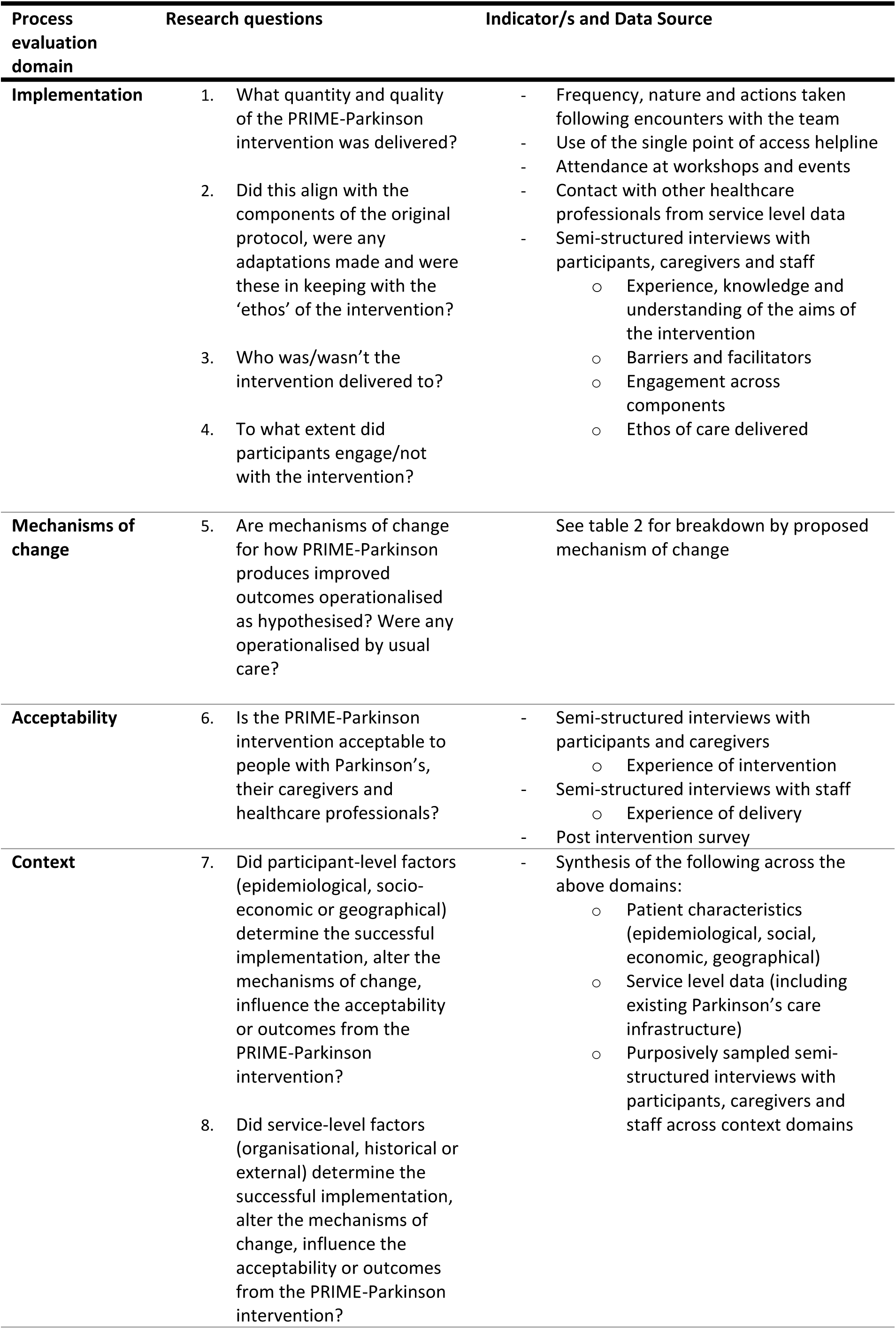
Process evaluation research questions within the four domains and indicators/data sources used to answer them.

## METHODS AND ANALYSIS

### Study design and sample size

The design of the primary study is a single-centre RCT to determine whether PRIME-Parkinson care can improve goal attainment as well as measures across multiple domains of health-related quality of life and symptom burden compared to usual care.[16] The sample size is 214 people with parkinsonism. A mixed methods approach will be conducted for the process evaluation outlined in this protocol. Data collection in the main trial started in October 2022 and will end in July 2025. Quantitative and qualitative data will be collected alongside trial outcome data between these time points to answer the specific process evaluation research questions.

All participants will contribute quantitative process evaluation data which will be collected prospectively throughout the study. Qualitative data for the process evaluation will be collected as part of a qualitative sub-study (“PRIME-Qual” IRAS 293614) from people with parkinsonism and caregivers enrolled in the main trial as well as members of the PRIME multidisciplinary team delivering the intervention and staff delivering ‘usual care’. We will recruit a purposive sample of around 10% recruited to the PRIME-UK RCT. This amounts to circa 10-15 patients and 10-15 caregivers from the active arm and the same from the control arm. We aim to achieve a varied sample in terms of variables which are anticipated to influence delivery and/or effectiveness of the intervention. Successful recruitment will be categorised as a sample containing:

- both male and female participants
- participants from each quartile of age from the study population
- a spread across time since diagnoses capturing those with early PD (diagnosed <2 years ago) and those living with PD for >10 years
- participants from across all 5 Hoehn and Yahr stages
- participants within low, medium and high risk of admission categories
- participants across all 3 local integrated care boards (ICBs)
- at least 1 participant from a nursing home
- participants that have and do not have an enrolled caregiver
- participants living in both rural and urban areas

We also aim to recruit between 10-15 staff to the study from those who are involved in the delivery of the new model of care and those involved in delivery of usual care.

## Data Sources and Data Collection

### Quantitative

Quantitative data relevant to the process evaluation will be collected prospectively as part of the trial for all participants. We will review data at 12, 18 and 24 months to guide iterative refinement of the intervention. Data collected will be as follows:

#### Record of encounters with clinical team

Data on all encounters with the PRIME team will be collected prospectively including number and duration of appointment, setting (home visit, clinic, inpatient, telephone, email), nature of appointment (action planning meeting, patient activated, clinician activated) and healthcare professional seen (including career stage). Actions taken from the clinic will also be recorded on an action log. Documented actions will fit into the following categories determined a priori: referrals to other specialities and allied healthcare professionals (AHPs), medication changes made, investigations requested and patient information provided (direct signposting to a website or paper resource). Further categorisation may be performed retrospectively e.g. referrals by specialism or AHPs.

#### Use of single point of access helpline

We will record all incoming calls to the single point of access helpline. We will capture who the call was from, nature of call, outcome of triage and action from team. We will also calculate time taken to reply to queries.

#### Delivery of education workshops and attendance

We will record the number of educational sessions aimed at participants, which are delivered as part of the intervention including duration, topic and speakers. We will record attendance, including whether a caregiver joined the participant.

#### Questionnaire data

Data from the following questionnaires collected as part of the trial will be extracted for the process evaluation: Patient Activation Measure (PAM)[20], Patients Assessment Chronic Illness Care (PACIC)[21] and Perceived Social Support[22]. The PACIC will be analysed as an overall score, and within its 5 domains (patient activation, delivery system/practice design, goal setting/tailoring, problem solving/contextual and follow-up/coordination)[21]. An intervention specific post-intervention survey will also be administered to those participants in the active arm upon study completion. This survey (see supplemental materials) presents statements to participants with a Likert-type scale focusing on acceptability of the intervention and whether or not proposed mechanisms of change were activated as hypothesised. This is designed specifically with this intervention in mind, therefore is not previously validated and builds upon methodology used in similar studies.[23]

#### Patient characteristics

Baseline patient data from the trial will be extracted to support in assessing the influence of contextual factors. We will focus on characteristics that we hypothesise could be associated with differing intervention implementation, engagement, acceptability and mechanisms of change. Variables will include age, gender, stage of disease (Hoehn and Yahr stage), symptom burden (measures of motor (MDS-Unified Parkinson’s Disease Rating Scale [24]) and non-motor symptoms (MDS Non-Motor Rating Scale [25])), cognitive impairment (Montreal Cognitive Assessment [26]), frailty (Survey of Health, Ageing and Retirement in Europe-Frailty Index 75+[27]), location of home (rural, urban) or whether in residential/nursing home, availability of caregiver and care provision, life space mobility (LSA questionnaire [28]) and socioeconomic status (index of multiple deprivation decile from postcode).

#### Service level data

We will capture which speciality is providing care (Neurology, Geriatrics, Private healthcare) to participants, as well as the frequency of their usual care appointments. We will collect data on the availability of services within each commissioning region (integrated care board – ICB) in the study catchment area (Bath and North East Somerset (BaNES), Somerset, Wiltshire, South Gloucestershire). We will also collect data from services on whether ‘usual care’ has changed from before and after the Covid-19 pandemic to consider whether we are comparing to a different service, ‘usual care’ which is diminished by the effects of covid, rather than ‘usual care’. This comparison will require use of routinely collected data from Parkinson’s services on frequency of clinical review including allied healthcare professionals from pre-covid (2018-2019) compared with (2023-2024) for those in the control arm who had a diagnosis prior to 2018.

#### PRIME team activities and training

We will collect the frequency of and attendees at PRIME clinical team meetings, as well as training delivered to team members.

#### Use of acute services including non-elective hospital admissions

Hospital Episode Statistics, collected as part of the economic evaluation, will be used to examine whether use of acute services is less between the intervention and control arms as a measure of activation of this hypothesised mechanism of change.

#### Other healthcare contacts

For participants in the usual care and active arm, we will also use Hospital Episode Statistics collected as part of the economic evaluation to measure contact with Parkinson’s services including use of helplines, specialist review and contact with allied health care professionals to look for differences in care received between the intervention and usual care arms.

### Qualitative data

Qualitative data collection will take place as close to recruitment as possible to explore expectations and then at 12 and 24 months later in line with the RCT follow-up. Data collection from staff will take place at 24 months. Data will be collected as follows:

#### Interviews with participants

Semi-structured interviews will be conducted with patients and family members/caregivers from the intervention and usual care arms of the trial. If preferred, patients and family members/caregivers may be interviewed together if both parties agree. The interview will be conducted face to face either at the hospital or in the participant’s home, for patients and family members/caregivers, or their place of work for staff. If this is not possible, we will offer to conduct this either by telephone or via video call. Semi-structured interviews have been selected over focus groups or other qualitative approaches for several reasons. The communication challenges facing people with parkinsonism such as cognitive issues and speech disorders can make it difficult for some participants to contribute during group sessions. Especially given the intention to recruit a varied sample, covering the heterogeneity of characteristics in this population, it is important that the methodologies adopted support all participants to contribute. This may include the caregiver facilitating the person with parkinsonism to contribute, which may be a more challenging dynamic in a focus group setting.

We will design the topic guide based on the aims of the process evaluation (implementation, mechanism of change, acceptability and context) including the following domains:

### Intervention and usual care arms

- experiences of the care received in both the intervention and usual care arms
- how receiving the intervention or usual care may have influenced participant’s experience of living with parkinsonism
- how/whether intervention components impacted or influenced the care received drawing on hypothesised mechanisms of change from the logic model

`o` within control arm this will explore whether same/different mechanisms of change were activated compared to the intervention arm participants
- how acceptable participants found receiving the intervention or usual care
- whether organisational or personal factors influenced participant’s and staff’s experiences of the intervention or usual care Intervention arm only
- their perceived engagement with the intervention including barriers and facilitators
- what aspects of the intervention have been helpful or less helpful in terms of maintaining their physical, psychological and social wellbeing and their ability to manage their own disease

The topic guide will focus on generating data to address our research questions but will be flexible enough to allow us to explore issues raised by participants (for an outline of the topic guide see supplementary materials 2 – this is subject to iterations as the trial progresses).

The purpose of collecting qualitative data from interviews with the usual care participants is to evaluate whether there was differential activation of the proposed mechanisms of change between the two trial arms. It also aims to capture the context of what ‘usual care’ includes. Together this data will support in interpretation of the main study findings and also in understanding scalability of the intervention.

#### Interviews with the PRIME team

Through semi-structured interviews with a sample of staff we will explore the experiences of delivering the intervention including barriers and facilitators of delivering the new model of care, acceptability and perceived contextual factors. Any adaptations made to the intervention and whether these fit within the intended ethos of the intervention will also be discussed.

### Case studies

Individual case studies, although not representative of the study population as a whole, can provide an in-depth picture of the intervention at the participant level. We will extract case studies of implementation of, and engagement with, the intervention to highlight how it was delivered in different scenarios, to demonstrate any mechanisms of change activated and highlight outcomes seen on a case-by-case basis. These cases will be purposively sampled by the study team to capture participants with high and low levels of engagement with the intervention. Case studies that illustrate activation of hypothesised mechanisms of change and situations where these were not will also be presented.

### Data Analysis

Quantitative data relevant to the process evaluation will be extracted from the main trial data and combined with qualitative analysis from the “PRIME-Qual” sub-study to form the process evaluation dataset. Analysis of the process evaluation data will be conducted separately to the study outcome data.

#### Quantitative

Quantitative data will be reported as descriptive statistics of observational data related to implementation (including engagement and fidelity), quantitative process measures related to mechanisms of change and whether this differs with measured contextual factors (organisational and personal characteristics) and between arms. In particular, within the active arm, we will identify groups in receipt of high/low levels of intervention and high/low levels of engagement and look for differences in characteristics. We will also examine whether high/low intervention/engagement groups having different activation of mechanisms of change.

We will combine process and outcome data to examine whether implementation (including engagement and fidelity) and activated mechanisms of change are associated with variation in trial outcomes using linear regression. We will also explore whether any identified relationships are moderated or mediated by contextual factors. This analysis will be predominantly exploratory in nature given the power will likely be inadequate.

#### Qualitative

The interviews will be audio recorded, transcribed and anonymised prior to analysis. The transcripts will be analysed thematically [19] using interpretive qualitative analysis involving induction and constant comparison. This will involve an iterative process of close reading of the data, coding, constant comparison and elaboration of emerging themes [20, 21]. In the first instance, two members of the research team will independently review a sample of three transcripts to develop and agree a coding strategy that reflects the process evaluation objectives. One researcher will then take responsibility for ongoing coding and categorisation of the data, using the QRS NVivo qualitative data management software. To assure the reliability of the coding and analysis process, codes and categories will be reviewed regularly by the wider team to ensure the accuracy of interpretation and internal consistency of codes. Categories of data and thematic relationships will then be identified and written up as descriptive and interpretive accounts, supported by interview excerpts.

### Combining quantitative and qualitative data

Measurement of the hypothesised mechanisms of change as detailed in the logic model will at times include the integration of quantitative and qualitative sources. A summary of how different data sources will be used as a marker of activation of each hypothesised mechanism of change is illustrated in Table 2.

**Table 2.** Mixed methods data sources to measure each proposed mechanism of change as per the logic model. Bold = quantitative, Italic = qualitative. PACIC = Patients Assessment Chronic Illness Care[21]

| Proposed Mechanism of Change | Data Source |
| --- | --- |
| Effective and personalised care and advice | **Record of encounters with clinical team**<br>**Use of single point of access helpline**<br>**Questionnaire (PACIC and post intervention survey)**<br>*Interviews with participants*<br>*Interviews with PRIME team*<br>*Case studies* |
| Increased ability to self-manage | **Questionnaires (Patient Activation Measure and post intervention survey)**<br>*Interviews with participants* |
| Rapid access to appropriate advice, review and signposting | **Record of encounters with clinical team**<br>**Use of single point of access helpline**<br>**Questionnaires (post-intervention survey)**<br>*Interviews with participants* |
| Minimising use or duration of use of acute services | **Use of acute services including non-elective hospital admissions**<br>**Questionnaires (post-intervention survey)** |
| Decreased feelings of loneliness, isolation, lack of specialist support | **Questionnaires (post-intervention survey)**<br>*Interviews with participants* |
| Empowerment to engage in own health promotion | **Delivery and engagement in education workshops**<br>**Questionnaires (Patient Activation Measure and post-intervention survey)**<br>*Interviews with participants* |
| Maintaining quality of intervention delivery | **Record of encounters with clinical team**<br>*Interview with PRIME team* |
| Improving efficacy and efficiency of multidisciplinary team | **PRIME team activities and training**<br>*Interviews with PRIME team* |

### Use of data to refine intervention

Process evaluation data on implementation and acceptability accumulated during the study will be used to guide refinement of the intervention. For example, if engagement with certain intervention components is poor, the interventional approach will be reviewed and adjusted to tackle deficiencies. Likewise, if only a certain patient group are receiving an intervention component, strategies may be employed to overcome this. Any refinements and adjustments made will be recorded and reported within the main results on effectiveness.

### Patient and public involvement

Guidance has been sought on the design of both the PRIME trial and the qualitative sub-study since study inception, including refining patient facing materials, study protocols and the consent process.[29] The was done through workshops and review of materials. We will be working with the patient public involvement (PPIE) group throughout the study to ensure that the intervention is designed with and for people who are affected by this condition and their caregivers.

## ETHICS AND DISSEMINATION

This process evaluation will integrate quantitative and qualitative data. The quantitative data will be collected as part of the main PRIME-UK RCT, which was granted NHS REC approval (21/LO/0387) on 27^th^ July 2021 and is registered on ClinicalTrials.gov (NCT05127057). The qualitative data will be collected as part of a sub-study, ‘PRIME-Qual’, which was granted NHS REC approval (21/LO/0388) on 14^th^ July 2021. The consent process for the main study is described in the original protocol and utilises a consultee system for those that lack capacity to consent to enrolment.[16] For ‘PRIME-Qual’, we will invite people with parkinsonism and their family members/caregivers who are recruited to the PRIME-UK RCT and members of the PRIME multidisciplinary team and usual care staff to take part. The inclusion/exclusion criteria are the same as those for the main trial.[16] Declining to participate in the ’PRIME Qual’ study will not preclude participation in the main RCT. Written consent will be sought from all participants and verbal assent will be sought at each subsequent interview. If it is not possible to collect written consent, then verbal consent will be taken and audio recorded. If participants lack capacity to consent, a personal consultee will be sought in accordance with the methods described in the original trial protocol.[16] We will invite all members of staff involved in delivery of the PRIME-Parkinson intervention to participate. They will be informed about the study via email, written information will be provided, and written consent will also be obtained by the research team. Findings of the process evaluation will be published in addition to the main trial findings.

## Data Availability

Access to the data will be available through application to the Chief Investigator. Pseudo-anonymised data may be shared with other researchers to enable knowledge synthesis.

## Author Contributions

KL: conception, study design, writing first draft, manuscript editing

EH/YBS: conception, supervision, refinement of methodology, manuscript editing

HB/SR: qualitative methodology, manuscript editing

SS: methodology, manuscript editing

ET/FL: main trial design, manuscript editing

JK: manuscript revisions with process evaluation expertise, manuscript editing

## Supplemental Materials 1 – Post intervention survey

This survey asks for your opinion on your experience of receiving PRIME care over the last 2 years and any impact it may have had on you or your care. The aim of this is to help us to better understand how, or not, the intervention works.

**For each of the statements, please circle the answer that best describes your experience.**

The results will be analysed anonymously, so please answer as honestly as possible.

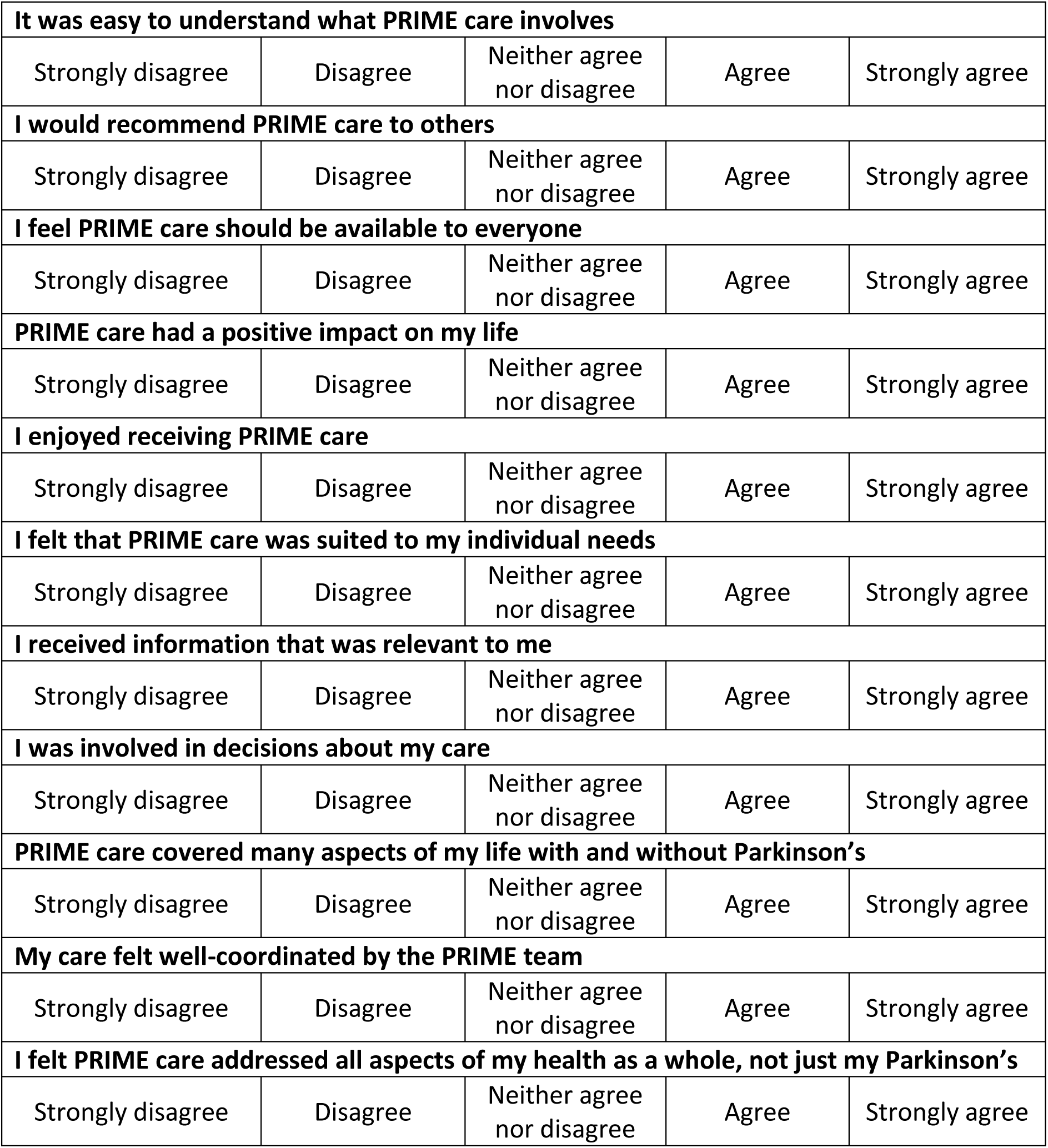
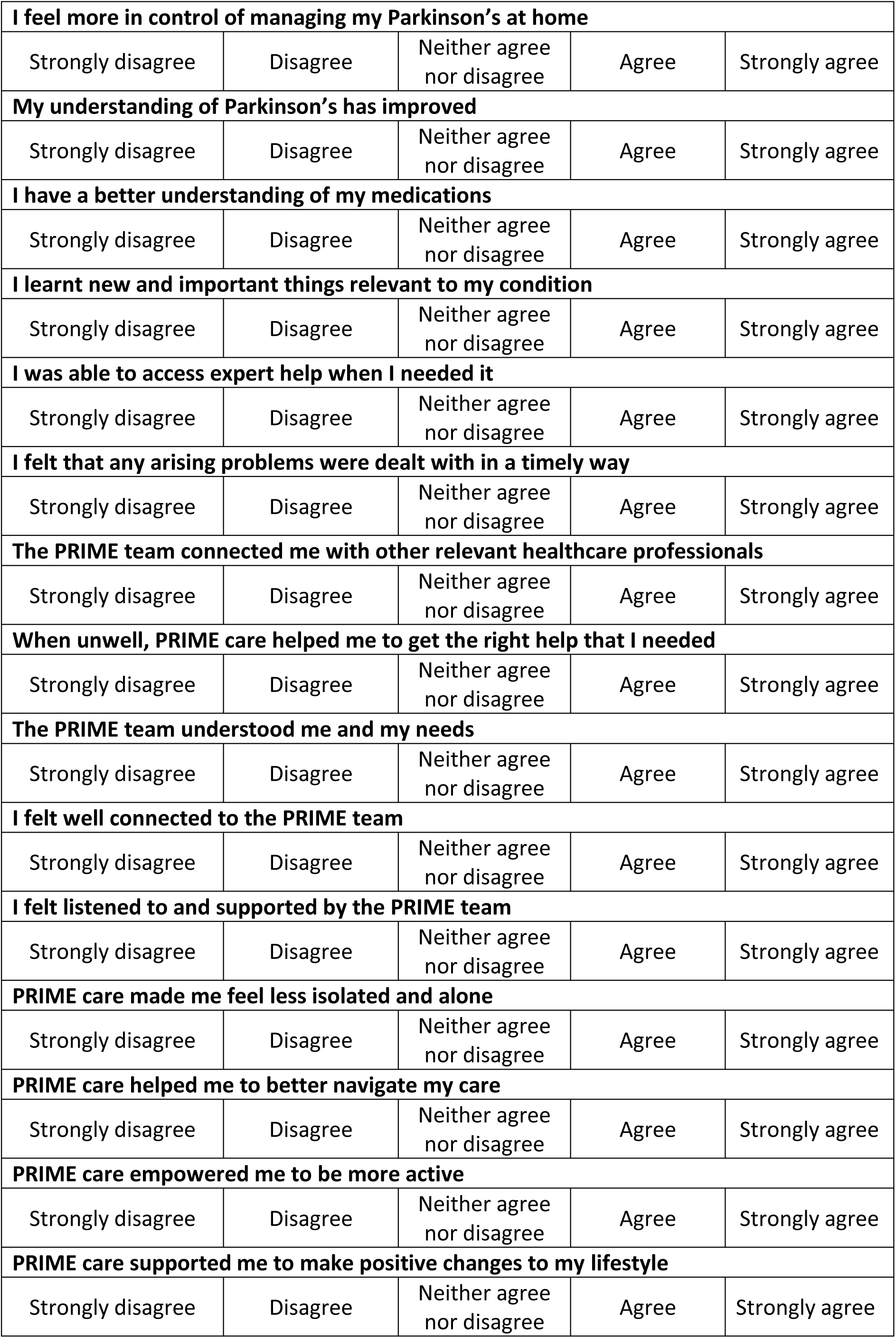

## Supplementary Materials 2 - Semi-structured interviews topic guide – PRIME-PD

For interviews with participants receiving **<u>PRIME care</u>**

**Introduction to the interview and aims:** Today’s interview will focus on your experience of Parkinson’s care including your experience of being in the PRIME study.

| OPENING QUESTIONS | Prompts |
| --- | --- |
| How have you been recently/since the last time you were interviewed for the PRIME study (reminder: by NAME, month and year of interview)? | Have there been any significant changes to your health/home situation/work (as applicable)? |
| What sort of things do you do to help you stay well? | [Physically, socially, emotionally]<br>This may relate to things you have learnt to do, medication, or therapies etc.<br>Can you give an example of something you/someone else does that helps?<br>Do you feel you have enough help and support? |
| Thinking about the care you receive in *general*, what has your experience of care been over the last 12 months? | What does your care consist of – who/what/where? [family members, professionals, aspects of care]<br>What aspects of this care are specific to PD? |
| Now thinking specifically about *PRIME* care: What do you understand about PRIME care? | What does it offer?<br>What does it aim to achieve? |
| What aspects of PRIME care have you received? | **Prompt as required:** your PRIME care might have included<br>- Assessments in clinic by the PRIME team<br>- The PRIME helpline<br>- PRIME workshops and groups<br>- A personal plan with the PRIME team |
| Have you seen the PRIME team in clinic for an assessment? | What happened during the assessment?<br>- Did the team assess your needs, make any changes, make onwards referrals?<br>What was your experience of it / How did you feel about this?<br>How knowledgeable did you find the staff? |
| Have you used the PRIME helpline number? | If yes, Tell me about your experience of the phone line<br>If you haven't used it, was there anything that stopped you from using it? |
| Have you attended any PRIME workshops or groups? | What was your experience of these?<br>Did anything encourage/stop you from attending these?<br>Did you learn anything from attending?<br>Did attending have any impact on how you live with PD? |
| Did you receive a personal plan for managing your PD from the PRIME team? | What did this involve? [discuss both how the plan was made and what actions it included]<br>How do you feel about the plan that was made? |
| What has your experience of the PRIME team been? | How often did you speak to the same person?<br>How well do you feel the PRIME team knew you and your situation? How did you feel about this? |
| Did anything make it easy to access/use/engage with PRIME care for you?<br>Was anything a barrier to you accessing/using/engaging with PRIME care? | Participant factors [stage of PD, cognitive impairment, relationship with existing cons, readiness to access care, socio-economic, having CG, access to transport, rural/urban, impact of covid] & Service factors [existing service infrastructure] |
| Thinking about your experiences of PRIME care overall, how do you feel about it? | What has been good or bad about PRIME care?<br>Can you give an example of a particularly good or bad experience of care?<br>Has it been more/less useful at different times over the last xx months? Why was this?<br>Has being in the trial had any other influences on you and your life? **Examples:** regular contacts, goal setting, being in a trial itself |
| Do you think the *PRIME* care you receive has impacted on your experience of living with PD or how you manage/approach your health? | Can you explain how/why this is the case? |
| Some people describe that their medical team doesn't really know them and what matters most – do you agree with this?<br>Has this changed since receiving PRIME care? | Did you feel that the PRIME team understood your needs?<br>Were you involved in making decisions about your care?<br>Did the care you received feel relevant to you and your priorities? |
| What is your experience of being able to access help when you need it? Was this different in PRIME care versus usual NHS care? | Do you feel you can get expert help in a timely way?<br>Can you think of a time you needed help for your PD and you could/could not access it? |
| Some people with PD describe feeling that they have a lack of specialist support, and this can leave them feeling like there's no one looking after them. What has your experience been?<br>Has this changed since receiving PRIME care? | What has your experience of the PRIME team been? |
| Did receiving PRIME care alter how/how much you contacted other health care professionals or visited the hospital? | How/why was this the case?<br>How did the PRIME team interact with other healthcare professionals? |
| How confident do you feel now about managing your PD yourself/taking control of your health? | What has led to this?<br>Has PRIME care impacted this?<br>**Prompts:**<br>- Do you feel you have more knowledge and understanding about PD?<br>- Have you gained information about PD? Where was this from? How easy was it to access?<br>- Do you feel more supported? By whom? How easy/difficult was it to access support?<br>Is there anything else that you think would help you feel more confident about managing your PD/health? |
| By taking part in this trial, you will also have had the chance to do some goal-setting. Can you tell me about your experience of this? | Did you find it changed how you behaved/acted/approached life?<br>Was it a negative or positive experience, and why? |
| As PRIME care comes to an end, how are you feeling about this? Do you have any concerns about this? | Are there any aspects you will miss?<br>Is there anything you will continue to do differently?<br>Do you think PRIME care differs from what you have received before, if so, how? |
| Overall, would you say your experience of PRIME was positive or negative? | Discuss aspects that were positive/negative<br>Was there anything you would suggest is changed or done differently?<br>Are there any aspects that should definitely be kept? |
| Would you recommend PRIME care to others? | What is it about PRIME care that means you would/wouldn't recommend this?<br>Do you think PRIME care should be available for everyone? |
| **CLOSING QUESTIONS** |  |
| Is there anything else you'd like to tell me about your experience of PRIME care and/or being part of the trial? |  |
| Do you have any questions you'd like to ask me? |  |

**Closing remarks:** Thank you for your time – if you have any questions please contact me. You will find my contact details at the end of the information booklet.

### Semi-structured interviews topic guide – PRIME-PD

For interviews with participants receiving **<u>usual care</u>**

**Introduction to the interview and aims:** Today’s interview will focus on your experience of Parkinson’s care including your experience of being in the PRIME study.

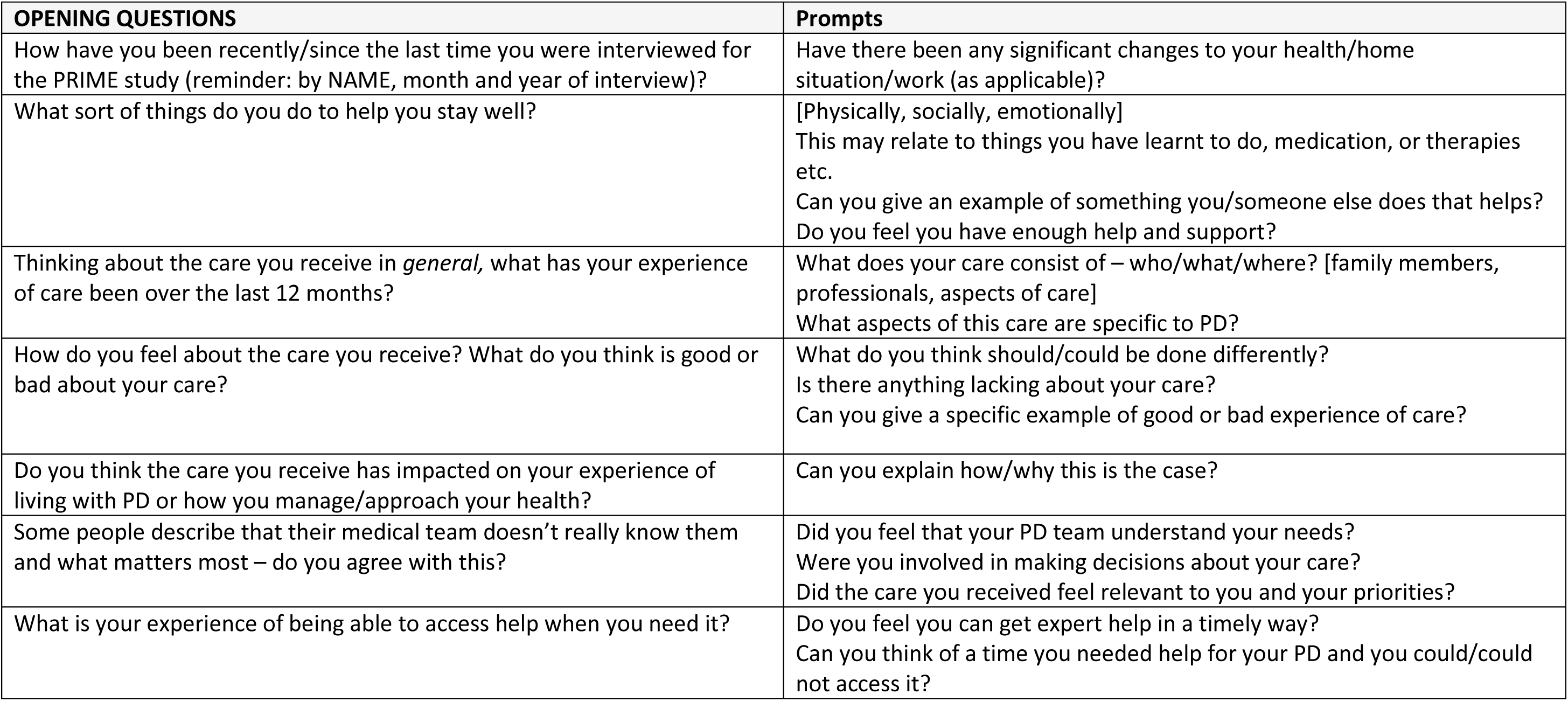
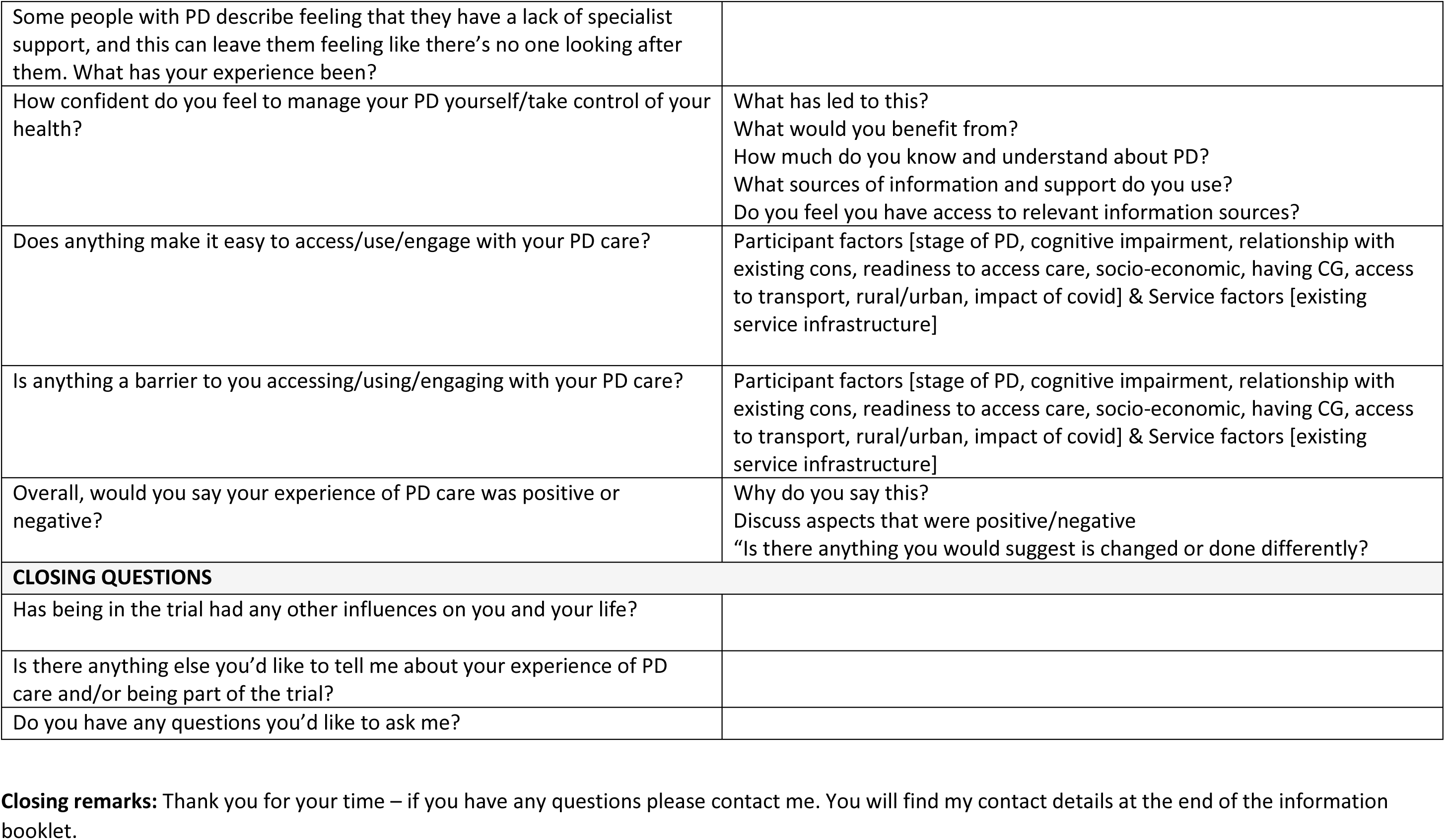

**Closing remarks:** Thank you for your time – if you have any questions please contact me. You will find my contact details at the end of the information booklet.

## REFERENCES

1 Ray Dorsey E, Elbaz A, Nichols E, et al. Global, regional, and national burden of Parkinson’s disease, 1990–2016: a systematic analysis for the Global Burden of Disease Study 2016. Lancet Neurol. 2018;17. doi: 10.1016/S1474-4422(18)30295-3

2 Feigin VL, Krishnamurthi R V., Theadom AM, et al. Global, regional, and national burden of neurological disorders during 1990–2015: a systematic analysis for the Global Burden of Disease Study 2015. Lancet Neurol. 2017;16. doi: 10.1016/S1474-4422(17)30299-5

3 Ishihara LS, Cheesbrough A, Brayne C, et al. Estimated life expectancy of Parkinson’s patients compared with the UK population. J Neurol Neurosurg Psychiatry. 2007;78. doi: 10.1136/jnnp.2006.100107

4 Schrag A, Jahanshahi M, Quinn N. How does Parkinson’s disease affect quality of life? A comparison with quality of life in the general population. Movement Disorders. 2000;15. doi: 10.1002/1531-8257(200011)15:6,1112::AID-MDS1008>3.0.CO;2-A

5 Schrag A, Hovris A, Morley D, et al. Caregiver-burden in parkinson’s disease is closely associated with psychiatric symptoms, falls, and disability. Parkinsonism Relat Disord. 2006;12. doi: 10.1016/j.parkreldis.2005.06.011

6 Huse DM, Schulman K, Orsini L, et al. Burden of illness in Parkinson’s disease. Movement Disorders. 2005;20. doi: 10.1002/mds.20609

7. Organization World Health. Delivering quality health services. 2018.

8 Greenland JC, Williams-Gray CH, Barker RA. The clinical heterogeneity of Parkinson’s disease and its therapeutic implications. European Journal of Neuroscience. 2019;49. doi: 10.1111/ejn.14094

9 Shulman LM, Taback RL, Rabinstein AA, et al. Non-recognition of depression and other non-motor symptoms in Parkinson’s disease. Parkinsonism Relat Disord. 2002;8. doi: 10.1016/S1353-8020(01)00015-3

10. Parkinson’s UK. UK Parkinson’s Audit. 2022.

11 Roberts AC, Rafferty MR, Wu SS, et al. Patterns and predictors of referrals to allied health services for individuals with Parkinson’s disease: A Parkinson’s foundation (PF) QII study. Parkinsonism Relat Disord. 2021;83:115–22. doi: 10.1016/J.PARKRELDIS.2020.11.024

12 Vlaanderen FP, Rompen L, Munneke M, et al. The voice of the Parkinson customer. J Parkinsons Dis. 2019;9. doi: 10.3233/JPD-181431

13 Bloem BR, Henderson EJ, Dorsey ER, et al. Integrated and patient-centred management of Parkinson’s disease: a network model for reshaping chronic neurological care. Lancet Neurol. 2020;19.

14 Tenison E, Smink A, Redwood S, et al. Proactive and Integrated Management and Empowerment in Parkinson’s Disease: Designing a New Model of Care. Parkinsons Dis. 2020;2020. doi: 10.1155/2020/8673087

15 Ypinga JHL, Van Halteren AD, Henderson EJ, et al. Rationale and design to evaluate the PRIME Parkinson care model: a prospective observational evaluation of proactive, integrated and patient-centred Parkinson care in The Netherlands (PRIME-NL). BMC Neurol. 2021;21. doi: 10.1186/s12883-021-02308-3

16 Lithander FE, Tenison E, Ypinga J, et al. Proactive and Integrated Management and Empowerment in Parkinson’s Disease protocol for a randomised controlled trial (PRIME-UK) to evaluate a new model of care. Trials. 2023;24. doi: 10.1186/s13063-023-07084-8

17 Skivington K, Matthews L, Simpson SA, et al. A new framework for developing and evaluating complex interventions: Update of Medical Research Council guidance. The BMJ. 2021;374. doi: 10.1136/bmj.n2061

18 Moore GF, Audrey S, Barker M, et al. Process evaluation of complex interventions: Medical Research Council guidance. BMJ (Online). 2015;350. doi: 10.1136/bmj.h1258

19 Whitehead M. A typology of actions to tackle social inequalities in health. J Epidemiol Community Health (1978). 2007;61. doi: 10.1136/jech.2005.037242

20 Hibbard JH, Mahoney ER, Stockard J, et al. Development and testing of a short form of the patient activation measure. Health Serv Res. 2005;40. doi: 10.1111/j.1475-6773.2005.00438.x

21 Glasgow RE, Wagner EH, Schaefer J, et al. Development and validation of the Patient Assessment of Chronic Illness Care (PACIC). Med Care. 2005;43. doi: 10.1097/01.mlr.0000160375.47920.8c

22 Zimet GD, Powell SS, Farley GK, et al. Psychometric Characteristics of the Multidimensional Scale of Perceived Social Support. J Pers Assess. 1990;55. doi: 10.1080/00223891.1990.9674095

23 Agley L, Hartley P, Duffill D, et al. Digital Intervention Promoting Physical Activity in People Newly Diagnosed with Parkinson’s Disease: Feasibility and Acceptability of the Knowledge, Exercise-Efficacy and Participation (KEEP) Intervention. J Parkinsons Dis. 2024;14:1193–210. doi: 10.3233/JPD-240071

24 Goetz CG, Tilley BC, Shaftman SR, et al. Movement Disorder Society-Sponsored Revision of the Unified Parkinson’s Disease Rating Scale (MDS-UPDRS): Scale presentation and clinimetric testing results. Movement Disorders. 2008;23. doi: 10.1002/mds.22340

25 Chaudhuri KR, Schrag A, Weintraub D, et al. The movement disorder society nonmotor rating scale: Initial validation study. Movement Disorders. 2020;35. doi: 10.1002/mds.27862

26 Nasreddine ZS, Phillips NA, Bédirian V, et al. The Montreal Cognitive Assessment, MoCA: A brief screening tool for mild cognitive impairment. J Am Geriatr Soc. 2005;53. doi: 10.1111/j.1532-5415.2005.53221.x

27 Romero-Ortuno R, Soraghan C. A Frailty Instrument for primary care for those aged 75 years or more: Findings from the Survey of Health, Ageing and Retirement in Europe, a longitudinal population-based cohort study (SHARE-FI75+). BMJ Open. 2014;4. doi: 10.1136/bmjopen-2014-006645

28 Parker M, Baker PS, Allman RM. A Life-Space Approach to Functional Assessment of Mobility in the Elderly. J Gerontol Soc Work. 2002;35. doi: 10.1300/J083v35n04_04

29 Lithander FE, Tenison E, Jones DA, et al. Working with public contributors in Parkinson’s research: What were the changes, benefits and learnings? A critical reflection from the researcher and public contributor perspective. Health Expectations. 2024;27. doi: 10.1111/hex.13914

